## Supplementary material for "Evaluating the cost-effectiveness of polygenic risk score-stratified screening for abdominal aortic aneurysm"

**S1	Effective sample size calculation**

The effective sample size ($N_{eff}$) for a study of a binary trait is equivalent to the sample size as if in the same study the number of cases or controls would have been equal. If the number of cases and controls are known, the following formula may be used to obtain this quantity:

$N_{eff}=\frac{4}{1/cases+ 1/controls}$.

In the scenario where the number of cases and controls is not known, $N_{eff}$ may be imputed via the following formula^1^:

$\hat{N_{eff}}=\frac{\frac{4}{{\sigma^{2}}_{G}}-\beta^{2}}{{\sigma_{\beta}}^{2}}$,

where ${\sigma^{2}}_{G}$ is the variance of the genotypes, $\beta$is the log odds ratio of the effect allele and $\sigma_{\beta}$ is the standard error of the log odds standard error. In the case of no genotype-level data being available, hence ${\sigma^{2}}_{G}$ is now known, it may be obtained from the allele frequencies of a matching reference panel via

${\sigma^{2}}_{G} =2 AF \left( 1-AF \right)$,

where $AF$ is the allele frequency of the effect allele.

The effective sample size calculation becomes more complex in meta-analyses, as in that case different SNPs may have different numbers of cases/controls that were available at that particular locus, which then results in the sample size becoming a distribution. Analyses that involve weighted meta-analyses of genetically overlapping traits, the sample size calculation becomes even more difficult, as in that scenario we have to account for the fact that at each variant the aetiology may only be partially shared between the different traits. The formula in this scenario to obtain the total per-SNP effective sample size ($N_{total}$) becomes^2^:

$N_{total} =N_{primary} \left( 1-\pi\right)+ N_{meta}\pi\left( 1-\rho\right)$,

where $N_{primary}$ is the effective sample size of the target trait and $N_{meta}$ is the effective sample size of the naive fixed-effects meta analysis between the primary and adjunct traits. $\pi$ is the local FDR (lFDR) providing the evidence of heterogeneity between the primary and adjunct trait at each SNP, and $\rho$ is the overlap between the primary and adjunct studies.

Furthermore, the vast majority of SNPs are not expected to contribute to either traits, hence those SNP’s ${N_{total}}$ is going to be ~$N_{meta}$, as their lFDR will be ~1. Therefore, naively interpreting the summary association results from such weighted meta-analysis may lead to a greatly overestimated${N_{total}}$, because even in unrelated traits a meta-analysis of null SNPs will yield the full combined sample size. Therefore, a more conservative estimate of ${N_{total}}$may be obtained by restricting the sample size calculation to only highly associated SNPs. By considering only the 447 SNPs with a combined association p < 5 * 10^-16^, the average sample size in our study was thus found to be 312,458, in contrast to the naive estimate of 458,939, if all SNPs were considered.

**S2	Genome-wide association studies of the AAA and AAA-related phenotypes**

Only HapMap3 panel of SNPs were considered, which were then filtered to eliminate those variants that failed QC in any of the UKB batches, or had a minor allele count < 20, or had an imputation INFO < 0.3. Samples were filtered out if they were not in the *“white.British.ancestry”* subset, or were too closely related, or were identified as sex discordant, all as defined in the UKB documentation. Finally, samples were also removed if they were part of the MetaGRS^3^ study.

The UKB cohort was split into two non-overlapping subsets, one for model training and the other for testing. The training set included all individuals on the BILIVE study and the Interim release of the datasets to avoid the known biases arising to the selection of participants BiLEVE study^4^, and to avoid any overlaps between the Nelson et al summary data^5^ and our own test set. An additional 10,125 controls were removed from our analyses that were used by our collaborators at the University of Leicester for their own association study.

For the AAA GWAS we excluded individuals from the controls who were on anti-hypertensive or lipid lowering medications as well as anyone who was in the AAA Related case list. This process yielded 1,068 cases and 127,011 controls and 133,900 cases and 127,011 controls for the AAA and AAA Related association studies, respectively. In turn, our AAA test set consisted of 869 cases and 91,012 controls, including 730 incident cases up until the age of 80.

For the genetic association step in our training set, we defined cases as those individuals who manifested the condition at any time within our records (as either prevalent or incident cases). For the model fit we also added in the following covariates: *age*, *sex*, *batch*chip* and the first ten principal components of ancestry. Association between phenotype and genotype was performed via PLINK2’s^6^ ‘*--glm firth-fallback*’ function.

**S3	Processing and quality control steps of association summary data**

The Malik et al^7^ and the AAAGen summary data lacked the per SNP breakdown of cases/controls, thus the effective sample size was imputed by the previously described method. Variants from all summary data were filtered to remove ambiguous SNPs (A/T and G/C), and those markers that were found to violate the following quality-control thresholds:

$$\sigma_{SS} <0.5 \sigma_{G} or \sigma_{SS} >\sigma_{G}+0.1 or \sigma_{SS} <0.1 or \sigma_{G} < 0.05 ,$$

where $\sigma_{SS}$ is defined as

$\sigma_{SS}=\frac{2}{\sqrt{{{{N_{eff}}_{\sigma}}_{\beta}}^{2}}}$.

The above threshold values used for this filtering step were sourced from the LDpred2 recommended settings^8^.

The summary data were combined via shaPRS^2^ in the following sequence, AAAGen, AAA in UKB, AAA Related, CAD and stroke. Note, we found that starting model training from the larger, better powered AAAGen study provided higher final accuracy results, as opposed to starting the AAA UKB study. We verified that the addition of each new summary data improved results at each stage. We also compared our results against combining all summary data in a single step via MTAG^9^, but found that combining the summary data via shaPRS performed better (an AUC of 0.699 vs 0.706, for MTAG vs shaPRS, respectively). Finally, we also compared PRS-CS against LDpred2 at fitting the best model, and found that LDpred2 provided a superior performance with an AUC of 0.707 vs 0.708 for PRS-CS vs LDpred2, respectively.

We ensured that our Test set evaluations were free from overfitting by working with a version of the AAAGen data that excluded the UKB. The full details of all PRS mode evaluation results can be found in Table S3.

**S4	Sensitivity analyses exploring impact of missing data**

We summarised missingness and explored the impact on results through the use of multiple imputation (MI) in a sensitivity analysis. Missing values were generated for risk factors using chained equations to create 20 imputed datasets with 10 burn-ins. Binary variables (alcohol intake, family history of CVD, diabetes, anti-hypertensive drugs, lipid-lowering drugs) were imputed using logistic regression, and smoking status using multinomial logit. Continuous variables (BMI, SBP, total cholesterol, HDL cholesterol) were imputed using predictive mean matching. The outcome indicator and cumulative hazard function (estimated with the Nelson-Aalen estimator) were included in all imputation models. Results from the 20 imputations were combined using Rubin’s rules to provide final estimates and are provided in Table S6.

| **Supplementary Table S1 \| AAA study details** | | |
| --- | --- | --- |
| **Phenotype** | **N cases** | **N Controls** |
| *UKB AAA* | 1,068 | 127,011 |
| *UKB AAA-related* | 133,900 | 127,011 |
| *AAAGen* | ~104,179.4 | |
| The number for cases/controls specified is the maximum per each study. Note, for the AAAGen dataset the number of cases/controls was unavailable, therefore the per-SNP effective sample size was imputed by the previously described methods. Thus the number quoted in the table is the median effective sample size across all SNPs. UKB AAA-related is a composite phenotype of conditions potentially genetically overlapping with AAA identified from the literature. | | |

| **Supplementary Table S2 \| UK Biobank Phenotype definitions used in PRS development** | |
| --- | --- |
| **Phenotype** | **ICD10 and OPSC4 codes** |
| *AAA* | **fatal_icd10:** I71.3 (Abdominal aortic aneurysm, ruptured), I71.4 (Abdominal aortic aneurysm, without rupture)  **nonfatal_icd10:** I71.3 (Abdominal aortic aneurysm, ruptured), I71.4 (Abdominal aortic aneurysm, without rupture)  **opcs4:** L18.3 (Emergency suprarenal open repair of AAA), L18.4 (Emergency infra-renal open repair of AAA), L18.5 (Emergency open repair of AAA (other)), L18.6 (Emergency infra-renal open repair of AAA (bifurcated)), L19.3 (Elective suprarenal open repair of AAA), L19.4 (Elective infra-renal open repair of AAA (straight graft)), L19.5 (Elective infra-renal open repair of AAA (other)), L19.6 (Elective infra-renal open repair of AAA (bifurcated  graft), L27.1 (Infra-renal EVAR), L27.2 (Suprarenal EVAR), L27.5 (EVAR at bifurcation NEC), L27.6 (Monoiliac EVAR), L28.1 (Infra-renal EVAR), L28.5 (EVAR at bifurcation NEC), L28.6 (Monoiliac EVAR) |
| *AAA-related* | **fatal_icd10:** Q87.4 (Marfan's syndrome), I21 (Acute myocardial infarction), I23 (Certain current complications following ST elevation (STEMI) and non-ST elevation (NSTEMI) myocardial infarction (within the 28 day period)), I22 (Subsequent ST elevation (STEMI) and non-ST elevation (NSTEMI) myocardial infarction), I25.2 (Old myocardial infarction), I70.2 (Atherosclerosis of native arteries of extremities with gangrene), I83 (Varicose veins of lower extremities), I86 (Varicose veins of other sites), Q79.6 (Ehlers-Danlos syndromes), M35.2 (Behçet's disease), M02.3 (Reiter's disease), M08.1 (Juvenile ankylosing spondylitis), M45 (Ankylosing spondylitis), E11 (Type 2 diabetes mellitus), M31.5 (Giant cell arteritis with polymyalgia rheumatica), M31.6 (Other giant cell arteritis), Q25.1 (Coarctation of aorta), Q61 (Cystic kidney disease), Q44.6 (Cystic disease of liver), E78 (Disorders of lipoprotein metabolism and other lipidemias), I10 (Essential (primary) hypertension)  **nonfatal_icd10:** same codes as fatal_icd10 |
| AAA phenotype is aortic abdominal aneurysm and AAA-related is a composite phenotype of conditions potentially genetically overlapping with AAA identified from the literature. | |

| **Supplementary Table S3 \| PRS model performance in test set** | | | | |
| --- | --- | --- | --- | --- |
| **Training dataset PRS is derived on** | **combined via** | **PRS** | **r^2^**  **(SD)** | **AUC**  **(LB -UB)** |
| *UKB AAA* | N/A | PRS-CS | 0.00034  (1.480*10^-5^) | 0.551  (0.532 - 0.570) |
| *AAAGen* |  |  | 0.00405  (1.480*10^-5^) | 0.682  (0.665 - 0.700) |
| *All AAA (starting from UKB)* | MTAG |  | 0.00444 (1.65*10^-5^) | 0.692 (0.675 - 0.709) |
|  | shaPRS |  | 0.00432 (1.65*10^-5^) | 0.689 (0.672 - 0.706) |
| *All AAA (starting from AAAGen)* | MTAG |  | 0.00317 (1.77*10^-5^) | 0.665 (0.647 - 0.683) |
|  | shaPRS |  | 0.00414 (1.71*10^-5^) | 0.684 (0.667 - 0.701) |
| *All AAA + AAA-related* | MTAG |  | 0.00509 (1.504*10^-5^) | 0.702 (0.685 - 0.719) |
|  | **shaPRS** |  | 0.00507 (1.53*10^-5^) | 0.704 (0.687 - 0.721) |
| ***All AAA + AAA-related + CAD*** |  |  | 0.00530 (1.66*10^-5^) | 0.707  (0.690 - 0.724) |
|  |  | **LDpred2** | **0.00547 (1.52*10^-5^)** | **0.708 (0.691 - 0.725)** |
| *All AAA + AAA-related + stroke* |  | PRS-CS | 0.00516 (1.36*10^-5^) | 0.705  (0.688 - 0.721) |
| *All AAA + AAA-related + CAD + stroke* | MTAG |  | 0.00497 (1.6*10^-5^) | 0.699  (0.682 - 0.716) |
|  | shaPRS |  | 0.00527 (1.53*10^-5^) | 0.706 (0.689 - 0.723) |
|  |  | LDpred2 | 0.00542 (1.45*10^-5^) | 0.707 (0.690 - 0.724) |
| Results for each of the datasets, PRS models and methods evaluated. The **data** column lists the studies that were used in the construction of the PRS. The **combined via** column displays the methods that were used to combine the association information from the studies. The **PRS** column contains the PRS construction methods that were used to generate the final PRS. The **r^2^** and **AUC** columns show the performance of the final PRS for each study calculated for differentiating AAA cases from controls in the test set. The r^2^ **SD** is calculated as the standard deviation of the squared correlation between the observed and predicted trait values resampled a 1,000 times. The AUC lower bound (**LB**) and upper bound (**UB**) represent the 95% confidence intervals which were computed with 2,000 stratified bootstrap replicates. Note, the AUC is calculated from a simple binary outcome including prevalent and incident events combined,with PRS included as a continuous variable with no other predictors included. | | | | |

| **Supplementary Table S4 \| Incident AAA code list used for time-to-event modelling** | |
| --- | --- |
| ICD codes | I71.3 (Abdominal aortic aneurysm, ruptured)*  I71.4 (Abdominal aortic aneurysm, without rupture)* |
| OPCS codes | L18.3 (Emergency suprarenal open repair of AAA), L18.4 (Emergency infra-renal open repair of AAA), L18.5 (Emergency open repair of AAA (other)) ,L18.6 (Emergency infra-renal open repair of AAA (bifurcated))  L19.3 (Elective suprarenal open repair of AAA), L19.4 (Elective infra-renal open repair of AAA (straight graft)), L19.5 (Elective infra-renal open repair of AAA (other)),L19.6 (Elective infra-renal open repair of AAA (bifurcated  graft))  L27.1 (Infra-renal EVAR), L27.2 (Suprarenal EVAR), L27.5 (EVAR at bifurcation NEC), L27.6 (Monoiliac EVAR)  L28.1 (Infra-renal EVAR), L28.2 (Suprarenal EVAR), L28.5 (EVAR at bifurcation NEC), L28.6 (Monoiliac EVAR) |

| **Supplementary Table S5 \| Summary of risk factors in UKB test set (N=91,731 with a PRS score)** | | | |
| --- | --- | --- | --- |
|  | **All** | **Males** | **Females** |
| N | 91,731 | 38,425 | 53,306 |
| Complete case N | 72,928 (79.5%) | 30,426 (79.2%) | 42,502 (79.7%) |
| Age at entry* | 56 (49, 62) | 56 (49, 62) | 57 (49, 62) |
| BMI (kg/m^2^)** | 26.6 (4.3) | 27.1 (3.8) | 26.2 (4.6) |
| Missing | 202 (0.2%) | 83 (0.2%) | 119 (0.2%) |
| Townsend index* | -2.5 (-3.8, -0.2) | -2.5 (-3.8, -0.1) | -2.5 (-3.8, -0.3) |
| Missing | 114 (0.1%) | 46 (0.1%) | 68 (0.1%) |
| Smoking status |  |  |  |
| Never | 37,038 (40%) | 13,528 (35%) | 23,510 (44%) |
| Ex | 46,520 (51%) | 20,695 (54%) | 25,825 (49%) |
| Current | 7,934 (9%) | 4,101 (11%) | 3833 (7%) |
| Missing | 239 (0.3%) | 101 (0.3%) | 138 (0.3%) |
| Any alcohol | 86,677 (95%) | 36,842 (96%) | 49,835 (93%) |
| Missing | 55 (<0.1%) | 23 (<0.1%) | 32 (<0.1%) |
| SBP (mm Hg)** | 134.9 (17.6) | 138.4 (16.3) | 132.4 (18.0) |
| Missing | 55 (<0.1%) | 11 (<0.1%) | 44 (<0.1%) |
| Anti-hypertensive drugs | 7,646 (8.4%) | 3,875 (10.1%) | 3,771 (7.1%) |
| Missing | 603 (0.7%) | 333 (0.9%) | 270 (0.5%) |
| Diabetes | 1,203 (1.3%) | 752 (2.0%) | 451 (0.9%) |
| Missing | 122 (0.2%) | 65 (0.2%) | 57 (0.1%) |
| Family history of CVD | 47,278 (56%) | 18,511 (53%) | 28,767 (58%) |
| Missing | 6,845 (7.5%) | 3,377 (8.8%) | 3,468 (6.5%) |
| Total cholesterol (mmol/L)** | 5.8 (1.1) | 5.7 (1.0) | 5.9 (1.1) |
| Missing | 4156 (4.5%) | 1698 (4.4%) | 2458 (4.6%) |
| HDL cholesterol (mmol/L)** | 1.5 (0.4) | 1.3 (0.3) | 1.6 (0.4) |
| Missing | 11,763 (12.8%) | 4,531 (11.8%) | 7,232 (13.6%) |
| Lipid lowering drugs | 7,292 (8.0%) | 4,087 (10.7%) | 3,205 (6.0%) |
| Missing | 603 (0.7%) | 333 (0.9%) | 270 (0.5%) |

* median (IQR)	** mean (SD)

Note: percentages given of those non-missing for each risk factor

| **Supplementary Table S6 \| Summary of observations by sub-group** | | | | |
| --- | --- | --- | --- | --- |
| **PRS tertile** | **Smoking status** | **Observed N** | **Observed proportion of population** | **Observed number of AAA events**  **(% of subgroup)*** |
| **Men** | | | | |
| All | All | 30,246 | 100% | 464 (1.53%) |
| Low | All | 10,770 | 35.4% | 58 (0.54%) |
| Intermediate |  | 9,908 | 32.6% | 136 (1.37%) |
| High |  | 9,748 | 32.0% | 270 (2.77%) |
| All | Never | 10,902 | 35.8% | 56 (0.51%) |
|  | Ex | 16,426 | 54.0% | 276 (1.68%) |
|  | Current | 3,098 | 10.2% | 132 (4.26%) |
| Low | Never | 3,960 | 13.0% | 16 (0.40%) |
| Intermediate |  | 3,537 | 11.6% | 17 (0.48%) |
| High |  | 3,405 | 11.2% | 23 (0.68%) |
| Low | Ex | 5,850 | 19.2% | 25 (0.43%) |
| Intermediate |  | 5,365 | 17.6% | 78 (1.45%) |
| High |  | 5,211 | 17.1% | 173 (3.32%) |
| Low | Current | 960 | 3.2% | 17 (1.77%) |
| Intermediate |  | 1,006 | 3.3% | 41 (4.08%) |
| High |  | 1,132 | 3.7% | 74 (6.54%) |
| **Women** | | | | |
| All | All | 42,502 | 100% | 81 (0.19%) |
| Low | All | 14,981 | 35.3% | 14 (0.09%) |
| Intermediate |  | 13,687 | 32.3% | 22 (0.16%) |
| High |  | 13,834 | 32.6% | 45 (0.33%) |
| All | Never | 18,941 | 44.6% | 17 (0.09%) |
|  | Ex | 20,652 | 48.6% | 41 (0.20%) |
|  | Current | 2,909 | 6.9% | 23 (0.79%) |
| Low | Never | 6,681 | 15.7% | 4 (0.06%) |
| Intermediate |  | 6,254 | 14.7% | 4 (0.06%) |
| High |  | 6,006 | 14.1% | 9 (0.15%) |
| Low | Ex | 7,433 | 17.5% | 5 (0.07%) |
| Intermediate |  | 6,503 | 15.3% | 11 (0.17%) |
| High |  | 6,716 | 15.8% | 25 (0.37%) |
| Low | Current | 867 | 2.0% | 5 (0.58%) |
| Intermediate |  | 930 | 2.2% | 7 (0.75%) |
| High |  | 1,112 | 2.6% | 11 (0.99%) |

* observed during whole follow-up period

| **Supplementary Table S7 \| Hazard ratios for recorded AAA from Cox regression, with multiply imputed dataset (N = 91,731)** | | |
| --- | --- | --- |
| **Risk factor** | **HR (95% CI)** | **p-value** |
| PRS group |  | <0.001 |
| Low risk | 1 |  |
| Intermediate risk | 2.33 (1.81, 3.01) |  |
| High risk | 4.46 (3.52, 5.66) |  |
| Sex |  | <0.001 |
| Female | 1 |  |
| Male | 4.77 (3.82, 5.95) |  |
| Townsend deprivation index (per 1 unit increase) | 1.02 (1.00, 1.05) | 0.05 |
| Alcohol intake |  | <0.001 |
| Non-drinker | 1 |  |
| Drinker | 0.56 (0.43, 0.72) |  |
| Family history of CVD |  | 0.6 |
| No | 1 |  |
| Yes | 1.04 (0.88, 1.22) |  |
| Diabetic |  | 0.09 |
| No | 1 |  |
| Yes | 1.26 (0.96, 1.65) |  |
| Smoking status |  | <0.001 |
| Never smoker | 1 |  |
| Ex-smoker | 2.39 (1.90, 2.99) |  |
| Current smoker | 8.07 (6.31, 10.33) |  |
| BMI  (per kg/m^2^ increase) | 1.01 (0.99, 1.03) | 0.5 |
| Systolic blood pressure  (per 10mm Hg) | 0.98 (0.94, 1.03) | 0.4 |
| Anti-hypertensive medication  No  Yes | 1  2.83 (2.38, 3.37) | <0.001 |
| Total cholesterol (per mmol/L) | 1.11 (1.03, 1.21) | 0.01 |
| HDL cholesterol (per mmol/L) | 0.28 (0.20, 0.38) | <0.001 |
| Lipid-lowering medication  No  Yes | 1  2.73 (2.26, 3.29) | <0.001 |

**Supplementary Figure 1**	Study Design

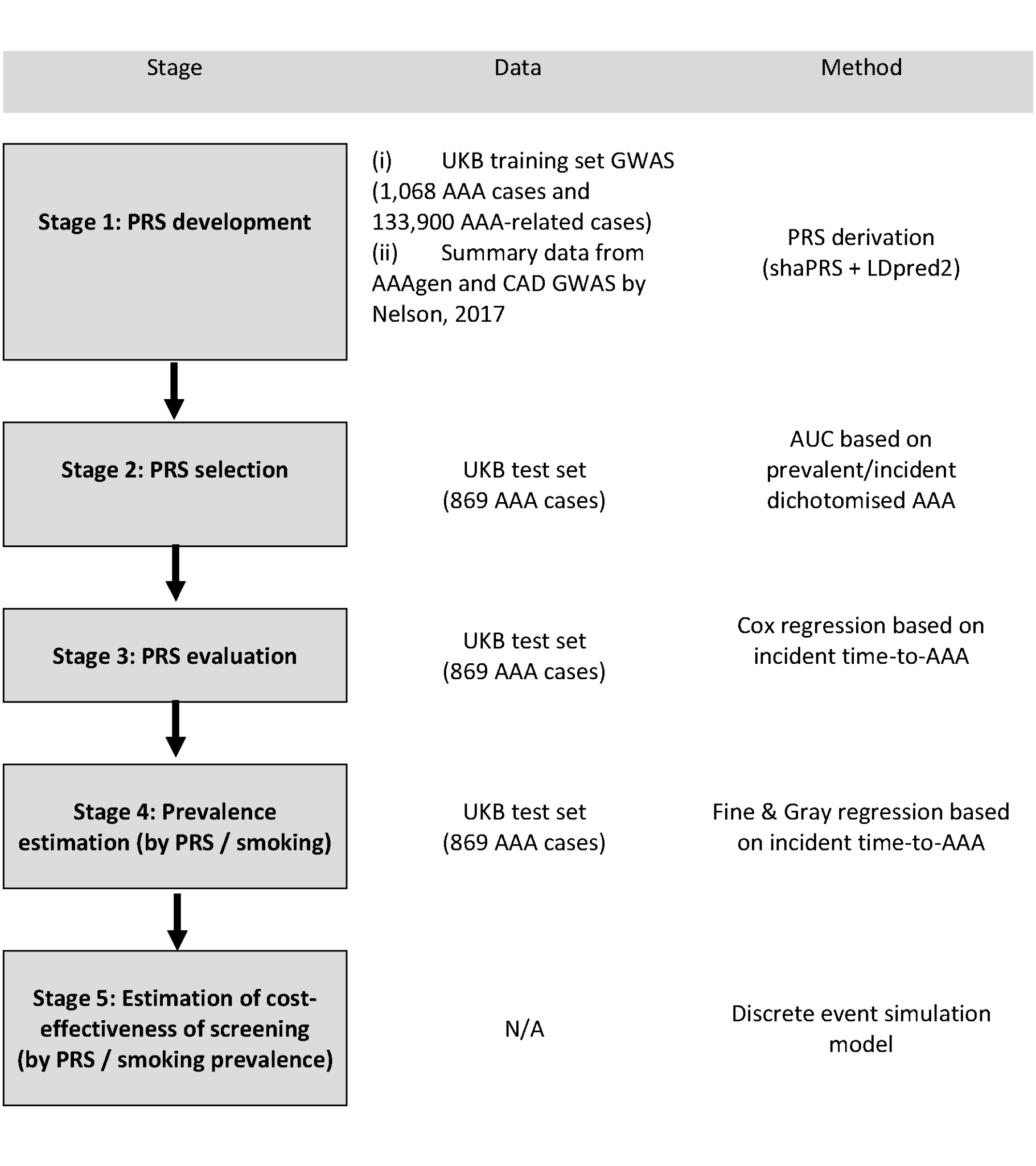

**Supplementary Figure 2**	Incremental net benefit compared to no invitation, by age at invitation and baseline prevalence at age 60 in men. INB is evaluated at a willingness-to-pay of £30,000 per QALY. Points plotted are point estimates with 95% uncertainty interval derived from 100 bootstrap PSA samples. Separate PRS and smoking sub-group prevalences estimated from UKB test set as CIF x inflation factor; indicated on the x-axis (PRS1 = low PRS risk group, PRS2 = intermediate PRS risk group, PRS3 = high PRS risk group; never = never smoker, ex = ex-smoker, curr = current smoker).

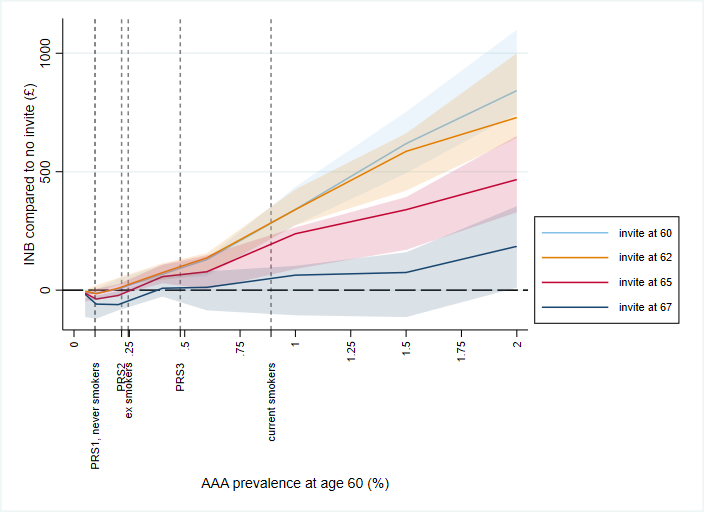

**Supplementary Figure 3**	Incremental net benefit compared to no invitation, by age at invitation and baseline prevalence at age 65 in women. INB is evaluated at a willingness-to-pay of £30,000 per QALY. Points plotted are point estimates with 95% uncertainty interval derived from 100 bootstrap PSA samples. Separate PRS and smoking sub-group prevalences estimated from UKB test set as CIF x inflation factor; indicated on the x-axis (PRS1 = low PRS risk group, PRS2 = intermediate PRS risk group, PRS3 = high PRS risk group; never = never smoker, ex = ex-smoker, curr = current smoker).

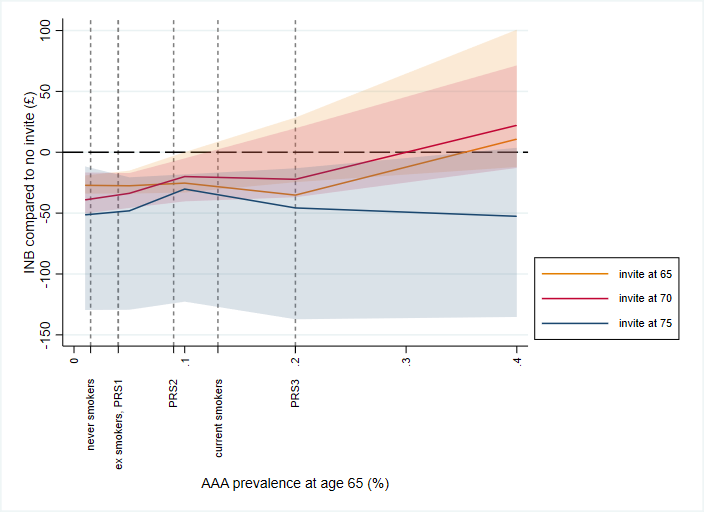

**Supplementary Figure 4**	Incremental net benefit compared to no invitation, by age at invitation and baseline prevalence at age 60 in men. INB is evaluated at a willingness-to-pay of £20,000 per QALY. Points plotted are point estimates with 95% uncertainty interval derived from 100 bootstrap PSA samples. PRS/smoking sub-group prevalences estimated from UKB test set as CIF x inflation factor; indicated on the x-axis (PRS1 = low PRS risk group, PRS2 = intermediate PRS risk group, PRS3 = high PRS risk group; never = never smoker, ex = ex-smoker, curr = current smoker).

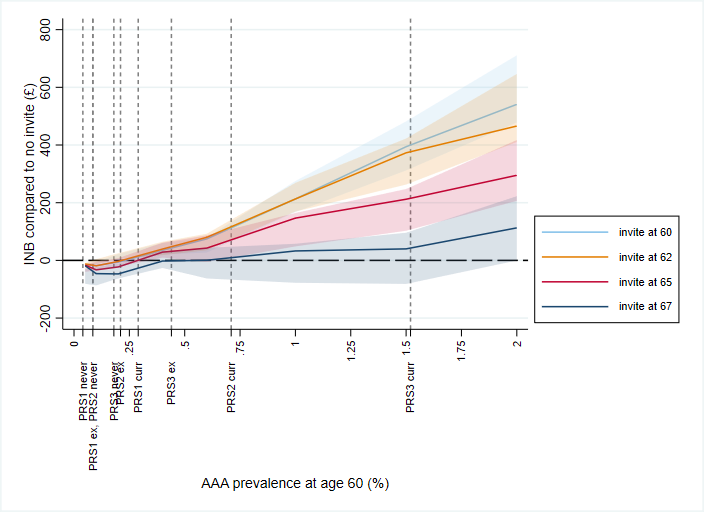

**Supplementary Figure 5**	Incremental net benefit compared to no invitation, by age at invitation and baseline prevalence at age 65 in women. INB is evaluated at a willingness-to-pay of £20,000 per QALY. Points plotted are point estimates with 95% uncertainty interval derived from 100 bootstrap PSA samples. PRS/smoking sub-group prevalences estimated from UKB test set as CIF x inflation factor; indicated on the x-axis (PRS1 = low PRS risk group, PRS2 = intermediate PRS risk group, PRS3 = high PRS risk group; never = never smoker, ex = ex-smoker, curr = current smoker).

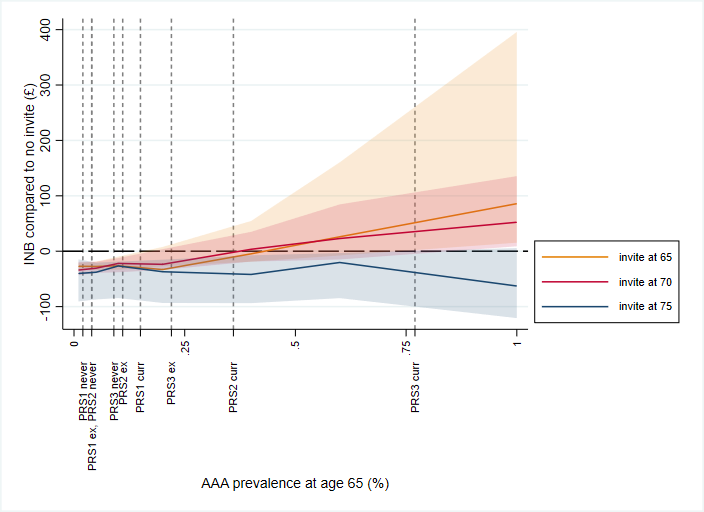
